# Genomic correlates with survival after treatment of oesophageal adenocarcinoma by surgery alone

**DOI:** 10.64898/2026.10.14.26365576

**Authors:** Anne Trinh, Thejaani P. Udumanne, Sarah J. Lord, Dan Falkenback, Angelique J. Levert-Mignon, Yi Jin Liew, Dana Pascovici, Michael J. Bourke, John P. Grady, Mark McCabe, Marjan M. Naeini, Anthony Joshua, Megan Barnet, Leonard D. Goldstein, Jason Ross, Jan Johansson, Reginald V. Lord

**Affiliations:** Data Science Platform, Garvan Institute of Medical Research, Sydney Australia; School of Clinical Medicine, St Vincent’s Healthcare Clinical Campus, UNSW Medicine & Health, UNSW Sydney, Sydney Australia; Gastroesophageal Cancer Research Program, St. Vincent’s Centre for Applied Medical Research, Sydney Australia; Sydney Clinical Trials Centre, University of Sydney, Sydney, Australia; Department of Surgery, Lund University, Lund, Sweden; CSIRO Health and Biosecurity, Westmead, Australia; Department of Gastrointestinal Endoscopy, Westmead Hospital, Sydney, Australia; Department of Medicine, University of Sydney, Sydney, Australia; St. Vincent’s Hospital, Sydney, Australia; Department of Surgery, University of Notre Dame School of Medicine, Sydney, Australia

**Keywords:** or phrases: oesophageal cancer, oesophageal adenocarcinoma, prognostic factors, genomic analysis, genomic sequencing

## Abstract

New prognostic biomarkers for oesophageal adenocarcinoma (OAC) may help treatment planning as current staging does not accurately predict survival for many patients. We used a 451 cancer-associated gene panel to assess genomic alterations associated with survival for a cohort of 65 OAC patients who underwent surgical resection without perioperative treatment, reflecting institutional practice at the time. Median age at operation was 72 years, 88% were male, median 32 lymph nodes were removed per patient, and minimum follow-up exceeded 11 years. TNM stages (pTNM, 8^th^ ed.) were 14% stage 1, 14% stage 2, 45% stage 3, and 6% stage 4 (non-locoregional nodal metastases). Copy number changes were common, and somatic mutations were similar to those reported previously. Genetic associations with worse survival included amplification of *CCND1* (HR=3.08, p=0.02), *EGFR* (HR=2.98, p=0.01), loss of *ARID2* (HR=2.12, p=0.05), *KMT2D* (HR=1.99, p=0.07) and missense mutation within *RYR1* (HR=2.68, p=0.01). This study identifies candidate genomic prognostic biomarkers in OAC using a cohort treated by surgery alone, enabling assessment of tumour biology independent of the confounding effects of peri-operative systemic therapy and radiotherapy. Several of the identified alterations are potentially targetable, supporting further investigation of genomic profiling to refine prognostication and guide treatment selection.

## INTRODUCTION

Survival for patients with adenocarcinoma of the oesophagus and oesophagogastric junction (OAC) remains one of the worst of all cancers with 15-25% five-year survival rates [1]. The incidence of OAC has increased dramatically over the last 50 years in many countries, including an approximately sixfold increase in areas of the USA [2]. Improving the poor survival outcomes requires methods for earlier detection and improved prognostication and treatment.

Apart from patients with early stage superficial OAC, who can be staged and usually cured by endoscopic resection, initial treatment decisions for patients with OAC are made using the clinical tumour node metastasis stage (cTNM) using imaging, endoscopy, and histopathological information. Unfortunately, studies that compare cTNM with pathological staging of surgery specimens (pTNM) suggest that cTNM staging for OAC can be surprisingly inaccurate [3,4]. For example, a systematic review that included 5,213 patients with cT2N0M0 oesophageal cancer who had multimodality staging and were treated with up-front surgery found that combined clinical tumour and nodal stage matched pathological stage in only 19% of cases [3]. Tumour downstaging to T1 was reported in 41%, which is significant as it suggests that many of these patients may have been adequately treated with endoscopic therapy rather than oesophagectomy. Conversely, 28% were upstaged to T3 or higher, and 34% upstaged from node negative to node positive disease. This under-staging is also highly significant as it may result in undertreatment [3]. Even pTNM staging, which is based on pathological examination of esophagectomy specimens, has limited accuracy as it does not capture the underlying biological heterogeneity. For example, despite having apparently low-risk (node negative) stage 2A OAC, approximately 50% of patients die within 5 years [4]. Validated prognostic biomarkers could identify these higher-risk patients for more intensive therapy.

Genomic profiling has demonstrated that OAC is characterised predominantly by chromosomal instability (CIN) rather than recurrent point mutations [5,6]. Common genetic events include *TP53* loss, which is thought to drive progression from Barrett’s oesophagus, amplification of receptor tyrosine kinase (RTK) pathway genes such as *ERBB2*, *EGFR* [7,8], and alterations in cell-cycle regulators including *CCND1* amplification and *CDKN2A* loss [6,9]. While these molecular features present opportunities for targeted therapies, identifying true prognostic biomarkers remains challenging because contemporary cohorts receive some form of peri-operative systemic treatment, and/or radiotherapy in some cases. In this setting, survival reflects both tumour biology and treatment response, making it difficult to distinguish biomarkers of therapeutic sensitivity from independent determinants of prognosis.

In this study we sought to identify genomic alterations associated with prognosis that could complement current pathological staging and improve risk stratification for patients with OAC. Although numerous candidate prognostic biomarkers have been reported, they have not been validated to the extent required to be incorporated into clinical practice. We analysed a unique cohort of fresh frozen OAC specimens from patients with more than 11 years’ follow-up. All patients were treated with the same *en bloc* oesophagectomy operation, which includes extensive nodal dissection allowing comprehensive pathological assessment for TNM staging. Importantly, due to institutional practice at the time, all patients were treated by surgery alone. This enabled assessment of tumour biology independent of peri-operative systemic therapy or radiotherapy, allowing genomic profiling to identify true prognostic biomarkers, rather than predictive biomarkers for treatment response.

## RESULTS

65 patients with oesophageal adenocarcinoma were included in the study cohort. All underwent *en bloc* oesophagectomy with extended lymphadenectomy between 2006 to 2011 and had fresh frozen tissue samples collected. The clinicopathological features of the cohort are summarised in Table 1 and Figure 1a. Median age at operation was 72 years (range 48 to 87), 57 (88%) were male, and 36 (55%) had Siewert II (junctional) oesophagogastric cancer. At surgery, the median number of lymph nodes removed was 32 (range 17 to 72). Pathology-defined TNM stages (pTNM, 8^th^ ed.) were stage 1: 9 patients (14%); stage 2: 9 patients (14%); stage 3: 29 patients (45%); and stage 4: 18 patients (28%) (Table 1).

**Figure 1:**
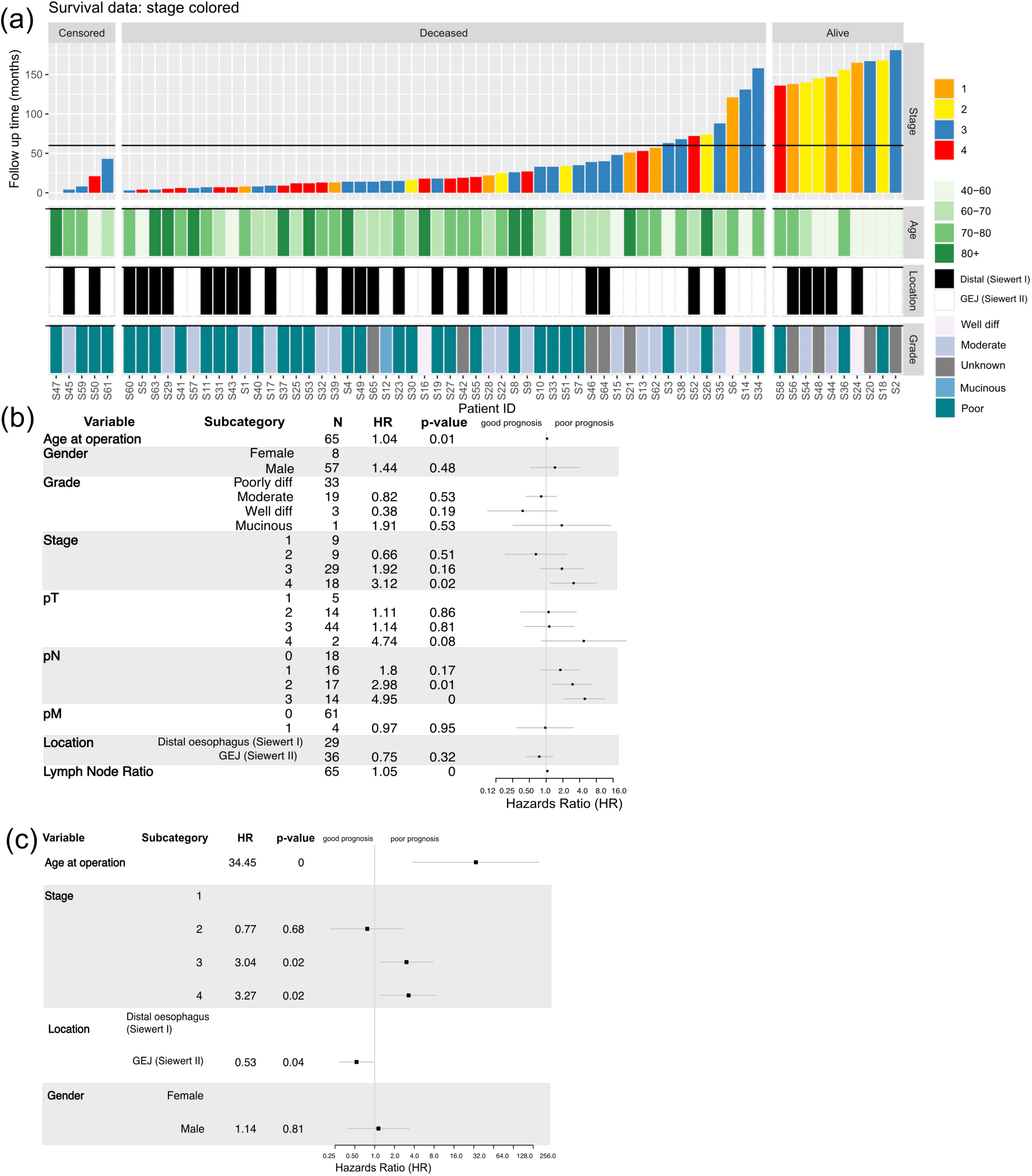
The clinicopathological features of the cohort. **a** Bar plot of follow up after surgery to death. Horizontal line below indicates a follow-up of 5 years. Five patients were censored due to non-cancer related death **b** Association of clinical characteristics with outcome using univariate cox proportional hazards model **c** Multivariate cox proportional hazards model for outcome including age, sex, stage and location.

**Table 1.**
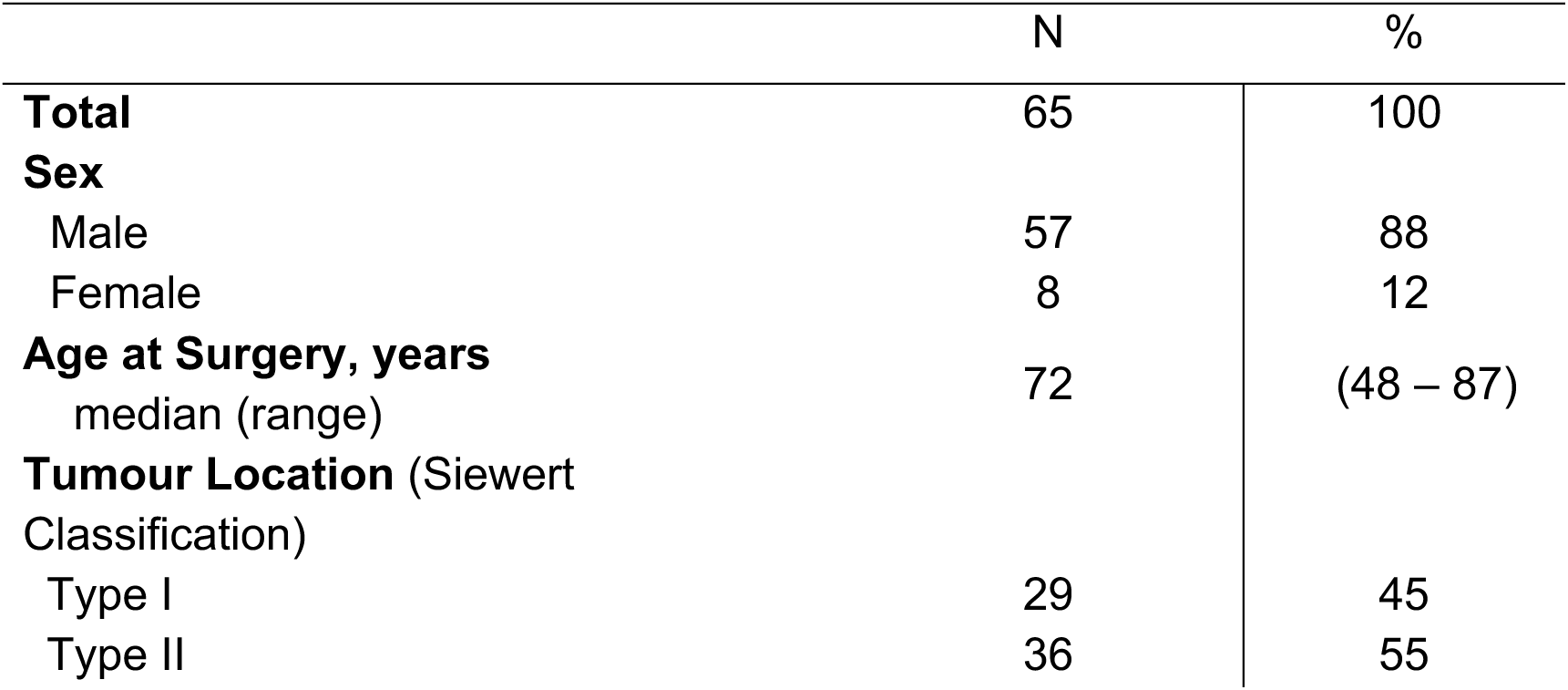
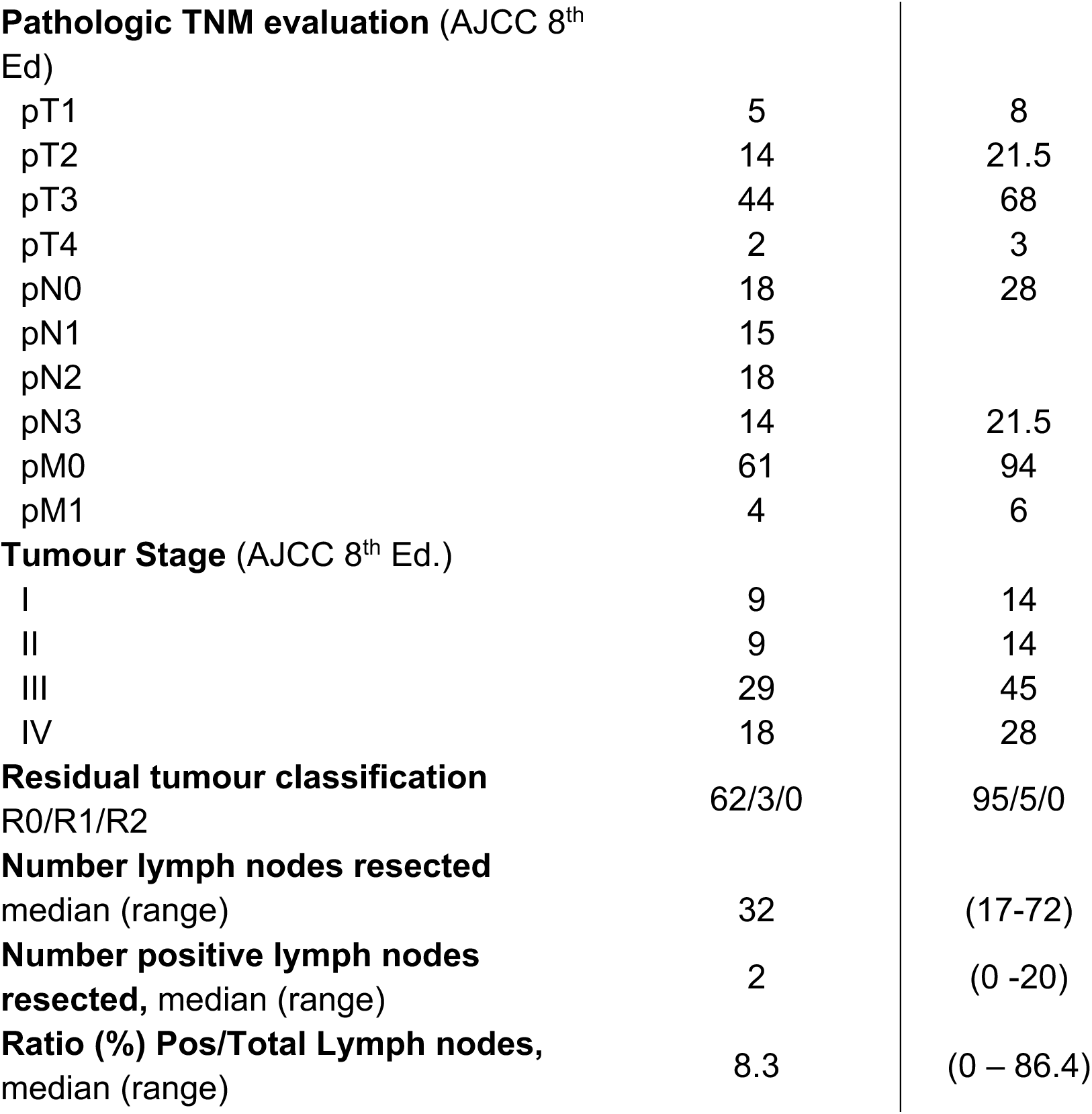
Clinical and pathological characteristics, patients with oesophageal adenocarcinoma and en bloc oesophagectomy, Lund University, Sweden 2006-2011.

### 11 year follow-up after en bloc oesophagectomy

At final follow-up at July 2022, 10 (15%) patients remained alive and had a minimum of 11 years follow-up (range 11 years, 4 months – 15 years, 1 month, Figure 1a). Median overall survival was 22 months (range 0-181 months, IQR 12 - 68 months). Twelve patients (18%) remained alive at 10 years, including five patients with advanced disease at diagnosis (four with Stage 3 and one with Stage 4 disease). In univariate Cox proportional hazards models, each additional year of age was associated with a 4% higher hazard of all-cause death, and stage 4 cancers were associated with a 3.1-fold higher hazard of all-cause death relative to stage 1 (Figure 1b). The strength of association between stage and overall survival remained similar in a multivariable model adjusting for age, sex, and cancer location. Cancer located in the distal oesophagus (Siewert I) was associated with a higher hazard of death than cancer considered to be arising from the oesophagogastric junction (Siewert II, adjusted HR=0.53, p=0.04); Figure 1c).

### Common Genomic Alterations in OAC

64 of 65 cancer samples and 59 matched squamous oesophageal mucosa control tissues were successfully sequenced and passed minimal quality metrics. Genomic profiling highlighted *TP53* as the most frequent non-synonymous single-nucleotide variant, affecting 49 (76.6%) of patients (Figure 2a). The second most frequently mutated gene was *LRP1B* (40.6% of patients), followed by *RYR1* (20.3%), *SPTA1* (20.3%), *ARID2* (18.8%), *ARID1A* (17.2%) and *PIK3CA* (17.2%) (Figure 2a).

**Figure 2:**
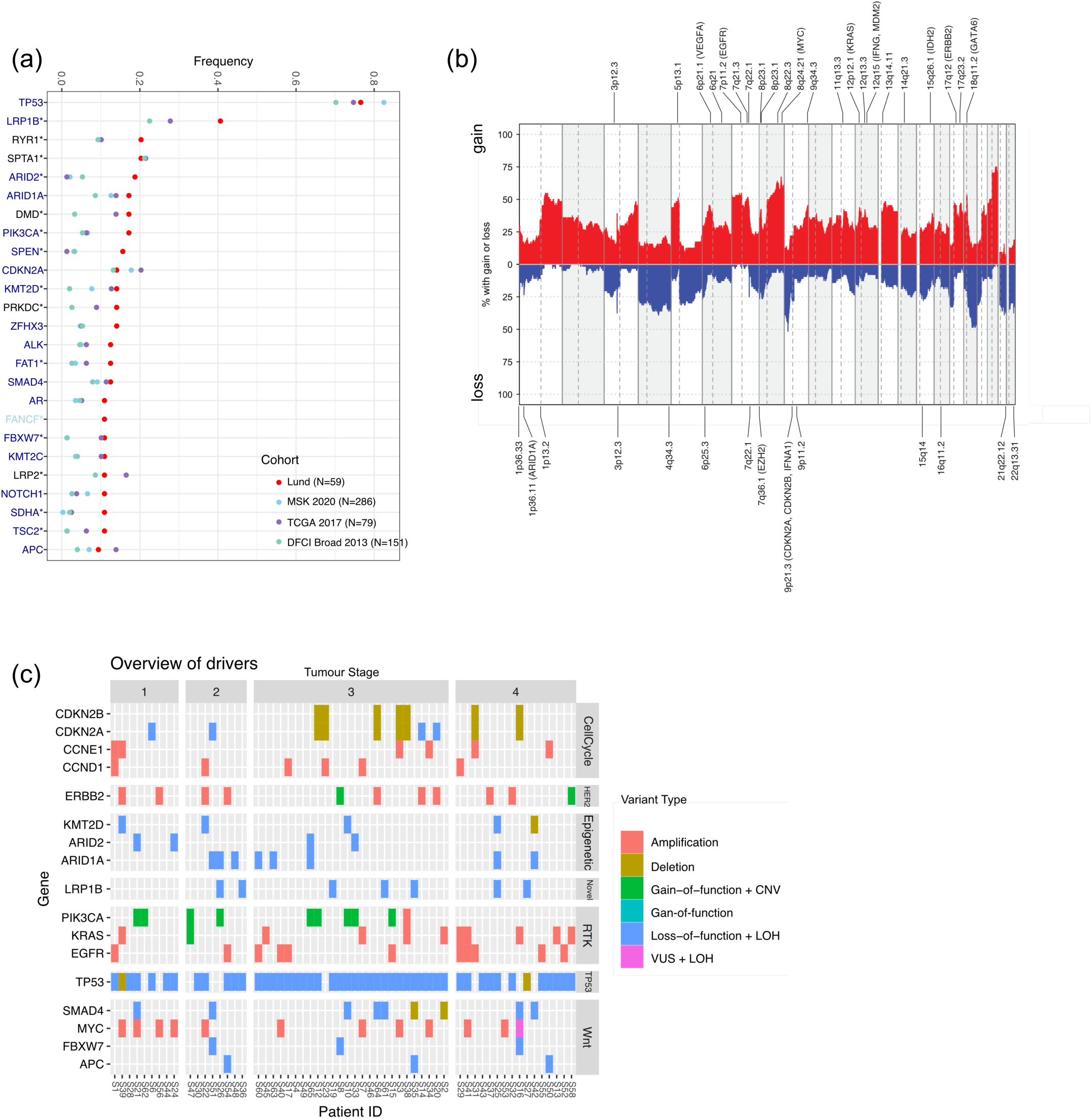
Genomic landscape of Lund OAC cohort. **a** Frequency of the 25 most commonly mutated genes in the Lund OAC cohort compared to the TCGA [7], MSK [8] and DFCI/Broad [5] studies. The significant genes have a p-value less than 0.05 based on a proportionality test (indicated with an asterisk) and thus appear at a different frequency compared to other public data. Gene label colour dark blue and light blue indicate driver event/COSMIC oesophageal specific genes and COSMIC cancer genes respectively. **b** Copy number alterations (defined log fold change of 0.3 due to low cellularity). Significant regions determined by GISTIC (qval<0.1). **c** Combined overview of SNV-CNV driver events in the cohort, stratified by tumour stage.

Compared to previously annotated cohorts from MSK [8], TCGA [7] and DFCI [5], *LRP1B*, *RYR1*, *ARID2* and *PIK3CA* mutations appeared at higher frequencies than expected, with two of these (*ARID2, PIK3CA)* predicted to be driver mutations [6]. Other predicted driver events in this cohort include mutations in *TP53*, *ARID1A*, *SPEN*, *CDK2NA* and *KMT2D*. These genes included those involved in nucleosome remodelling (*ARID1A*, *SMAD4*, *KMT2D*) and cell-cycle aberrations (*CDKN2A*), with cohort aberration frequencies less than 20%, which is similar to that observed in previously published cohorts [5,7,8] (Supplementary Figure 1).

Copy number profiling was relatively similar to previous studies, showing key amplifications in chromosomes 1q, 5p, 7p, 8q, 17q and deletions in 4, 9p. Notable common drivers on these chromosomes include oncogenes in the RTK signalling pathway, (*EGFR* (7p11, 17%), *KRAS* (12p, 15%), *ERBB2* (17q, 14%)), the proliferative-related gene *MYC* (8q, 17%) while 10% of patients showed co-deletion of *CDKN2A* and *CDKN2B* (9p) (Figure 2b) [7,9,10]. The full list of predicted driver events is presented in Supplementary Table 2.

### Commonly aberrated genomic pathways in OAC

Collapsing commonly aberrated genes at a combined mutational and CNV level illustrated four main aberrant pathways in this disease (Supplementary Fig 1, Figure 2c): 1. Cell-cycle regulation, 2. RTK-signalling, 3. Epigenetic modifications, and 4. Wnt signalling, aligning with previous genomic reports [5–8].

Cell-cycle regulation was characterised mainly by CNV events whereby 10% of patients showed co-deletion of *CDKN2A* and *CDKN2B* [7,9,10]; amplifications in *CCNE1* and *CCND1* were also common (Figure 2c). *TP53* loss of heterozygosity (LOH) through a point mutation and copy number loss was very common (Figure 2c). Alterations in epigenetic modifiers were predominantly loss of function (LoF) events (*KMT2D, ARID1A, ARID2*, Figure 2c), featuring genes involved in nucleosome remodelling (*ARID1A*, *SMAD4*, *KMT2D*).

The RTK signalling pathway is characterised mainly by chromosomal amplifications in *EGFR*, *KRAS* and *ERBB2. PIK3CA* was characterised by gain of function combined with amplification. Lastly, the Wnt signalling pathway aberrations included amplification in *MYC* and LOH events in genes including *SMAD4*, *FBXW7,* and *APC*.

### Genomic pathways associated with survival

First, we evaluated the association between the most frequently altered genes and overall survival, adjusting for age and pathological stage (Figure 3). These genes represented the four major pathways recurrently disrupted in OAC: cell-cycle regulation, epigenetic modification, RTK- and Wnt signalling. Across all stages, *CCND1* amplification was independently associated with worse survival (HR=3.08, p=0.02), whereas LoF or LOH alterations in *APC* were associated with improved survival (HR=0.24, p=0.03). In addition, aberrations in *RYR1*, not typically implicated in canonical OAC signalling pathways, were independently associated with poorer survival (HR=2.68, p=0.01). Of the 16 patients harbouring *RYR1* aberrations, 12 (20.3%) had SNV mutations and 5 (7.8%) had gene amplification. In contrast to other studies, *TP53* aberrations were not associated with survival.

**Figure 3:**
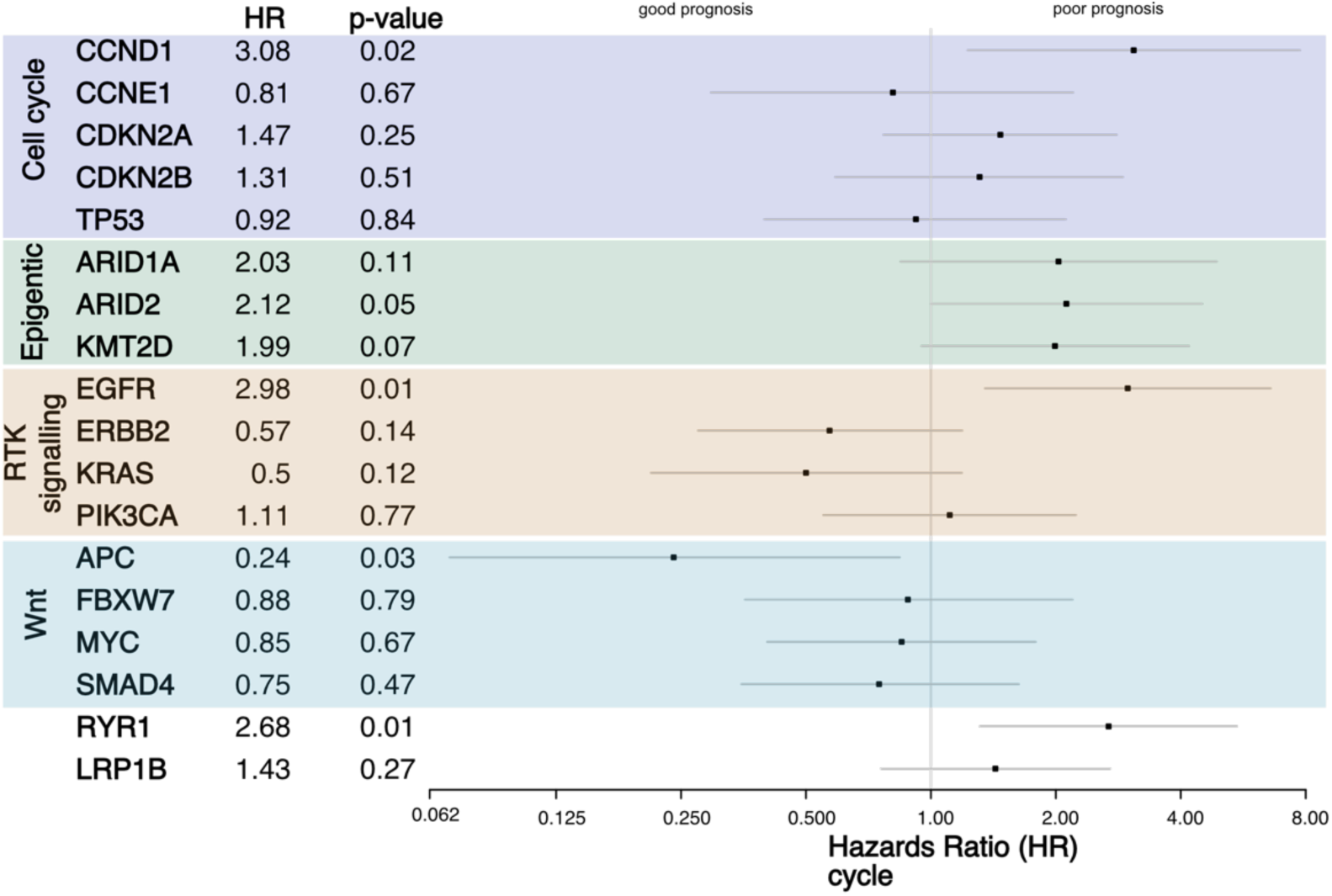
Association of variants with survival. Most frequent genes with SNV-CNVs within the cohort and associated hazards ratios of the variant using a univariate cox proportional hazards model adjusted to age and stage. Genes are grouped according to impacted pathway.

Within the epigenetic modifiers, LoF alterations in *ARID2* (HR=2.12, p=0.05) and *KMT2D* (HR=1.99, p=0.07) were independently associated with poorer survival. Within the RTK signalling pathway, contrasting associations with survival were observed. Activating alterations in *KRAS* and *ERBB2* (GoF mutations or amplification) were associated with improved survival (HR=0.5, p=0.12 and HR=0.57, p=0.14 respectively), whereas *EGFR* amplification was associated with poorer survival (HR=2.98, p=0.01).

### Tumour Mutation Burden (TMB) association with survival

As individual mutations and copy number changes appear at relatively low frequencies in OAC (<20%), we investigated whether the global measures of genomic instability tumour mutational burden (TMB) associated with survival. TMB was low overall (median ∼6 mutations/MB), however 5 patients had high TMB (> 10 mut/MB). One of these 5 patients harboured a somatic mutation in microsatellite gene *MSH2*, known to confer microsatellite instability. Four patients had *LRP1B* mutation and two had *RYR1* mutation; the large size of *LRP1B* and *RYR1* suggests that these genes could serve as surrogate markers for cumulative mutational burden. Indeed, stratification according to mutational status of *LRP1B* and *RYR1* demonstrated higher TMB compared to their wild-type counterparts (Figure 4a).

**Figure 4:**
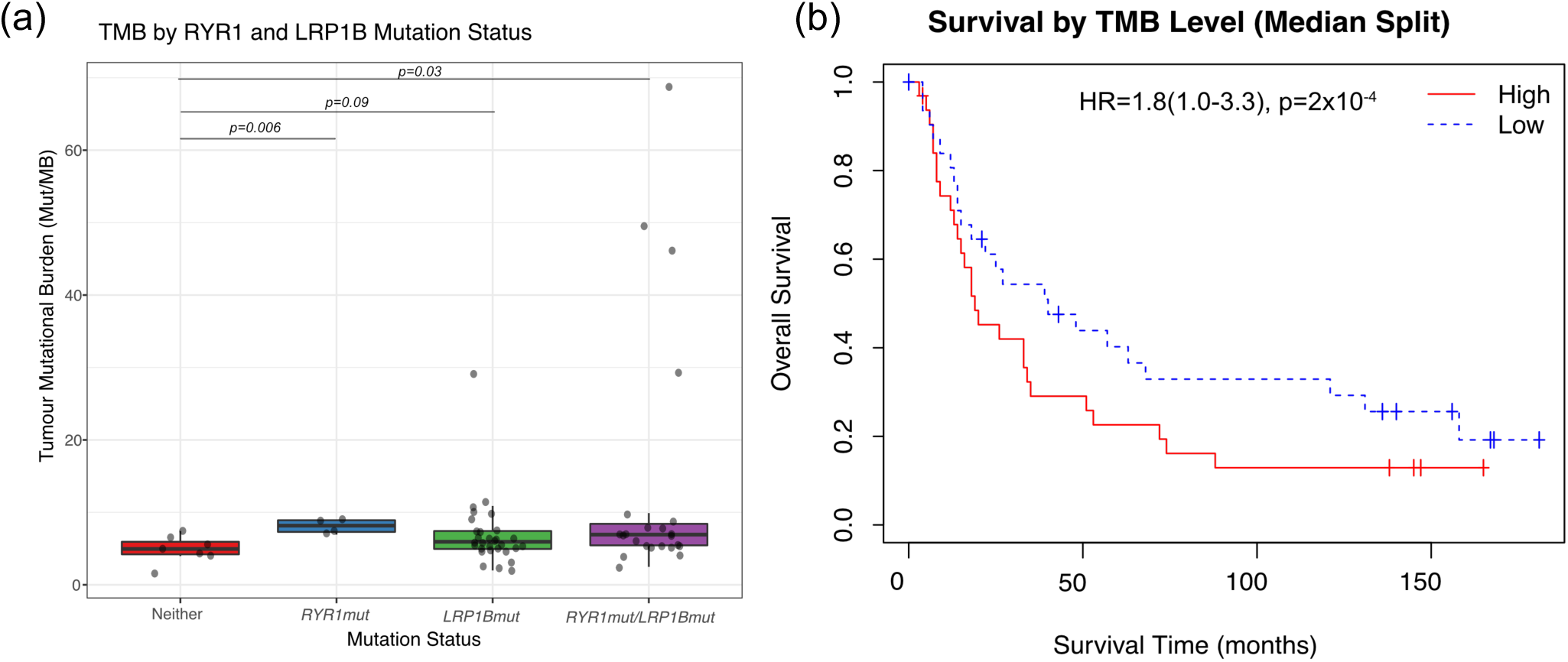
Association of samples with tumour mutational burden. **a** Tumour Mutational Burden in patient samples harbouring a *RYR1 and/or LRP1B* variants compared to those with wild type. Significance calculated **b** Kaplan Meier plot of survival in patients higher than vs below the median TMB. Hazards Ratio with 95% Confidence Interval, log-rank p test reported.

In a Cox proportional hazards model adjusted for age and pathological stage, patients with a TMB level below the cohort median had significantly improved survival (HR=0.55, 95% CI: 0.31-0.99, p=0.048, Figure 4b).

## DISCUSSION

This study reports genomic findings from a cohort of patients with adenocarcinoma of the oesophagus or oesophagogastric junction with more than 11 years follow-up. All patients underwent a standardised, extensive *en bloc* oesophagectomy without peri-operative therapy, despite most having stage 3 or 4 disease (the latter due to non-locoregional lymph node metastasis). Under current standards of care, fit patients would receive peri-operative chemotherapy with immunotherapy [11]. The absence of systemic or radiotherapy in this cohort provides a rare opportunity to examine the natural biological history of OAC. Consequently, genomic associations with survival identified in this study are more likely to represent prognostic biomarkers reflecting intrinsic tumour biology, rather than predictive biomarkers of response or resistance to peri-operative therapy.

Our findings regarding the type and frequency of genomic alterations are broadly consistent with previous studies, with recurrent aberrations affecting four principle biological pathways: (1) Cell-cycle regulation [5–7,10], (2) RTK-signalling [6,8], (3) Epigenetic modifiers [6,7,10], and (4) Wnt signalling [6]. Despite the relatively small cohort, several genomic alterations were significantly associated with overall survival after adjustment for age and pathological stage. In particular, aberrations affecting cell-cycle regulation and epigenetic remodelling were associated with worse prognosis, whereas contrasting survival associations were observed within the RTK signalling pathway, with activation or amplification of EGFR and KRAS/ERBB2 exhibiting opposing effects. No consistent prognostic associations were observed for the other pathways. Contrary to other studies [12], loss of function mutations or deletion of tumour suppressor *TP53* did not correlate with survival in our cohort.

The Cyclin D1-CDK4/6 complex promotes cell cycle progression from G1 to S phase by phosphorylation and inactivation of the retinoblastoma protein (Rb), resulting in the expression of S-phase specific genes involved in DNA replication and chromatin assembly. *CCND1* gene is frequently amplified in a variety of cancers, including OAC, in which it has been reported to be associated with worse prognosis and overall poor survival rates [13–15]. Consistent with these reports, *CCND1* amplification was independently associated with worse survival in our cohort. CDK4/6 inhibitors block phosphorylation of Rb, thereby preventing G1/S cell-cycle transition and suppressing tumour proliferation. Although this represents a rational therapeutic strategy in CCND1-amplified tumours, clinical activity in OAC has been modest. Single agent palbociclib showed limited efficacy [16], while combination therapy with abemaciclib (CDK4/6 inhibitor) and ramucirumab (VEGFR2 inhibitor) has shown some encouraging early activity, suggesting this strategy may benefit molecularly-selected patients [17].

To our knowledge, this is the first study to associate aberrations in epigenetic remodelling genes with poorer survival in OAC. *ARID1A* and *ARID2* are key components of SWI/SNF chromatin remodelling complex. Evidence suggests that *ARID1A* can act as either a tumour suppressor or an oncogene depending on the tumour type [18], whereas *ARID2* is primarily known for its role as a tumour suppressor and the presence of *ARID2* mutations is well-characterised in OAC [5,9]. Selective or co-mutation of these genes may result in epigenetic dysregulation that facilitates more aggressive clinical phenotypes.

The prognostic significance of *ARID1A* aberrations on survival remains uncertain, with studies reporting both favourable and adverse associations of *ARID1A* deficiency on survival [19–21]. Notably, although published analyses have excluded or stratified for patients according to receipt of neoadjuvant therapy, none has comprehensively annotated adjuvant treatment, limiting the ability to distinguish biomarkers of intrinsic prognosis from those predictive of treatment response. Beyond prognostic significance, *ARID1A*-deficient tumours have emerged as a potential therapeutic subgroup. An ongoing clinical trial (NCT06824363) is evaluating whether a single dose of dual immune checkpoint blockade can increase intra-tumoral T cell infiltrate in patients with *ARID1A*-mutant oesophageal, gastro-oesophageal junction, and gastric adenocarcinomas.

Histone methyltransferase KMT2D, a histone modifier involved in cellular development and differentiation, is frequently mutated and deleted in several cancers including oesophageal cancers, with more data available for oesophageal squamous cell carcinoma (ESCC) [22,23]. In our cohort, loss of *KMT2D* was associated with poorer survival, representing, to our knowledge, the first report linking *KMT2D* alterations to prognosis in OAC. Consistent with its role in maintaining epigenetic homeostasis, *KMT2D* is generally considered a tumour suppressor, including in ESCC [24], although context-dependent proto-oncogene function has been reported in gastric cancer [25]. Given recent advances in epigenetic therapies, including drugs specifically targeting other histone methyltransferases such as EZH2 and DOTL1, KMT2D may present a potential target for future OAC treatment strategies [26,27].

Epidermal growth factor receptor (*EGFR*) is a proto-oncogene that is often amplified in OAC [7,8]. Consistent with our findings, several studies have demonstrated that *EGFR* amplification and overexpression are associated with worse overall survival in patients with oesophageal and GOJ adenocarcinoma [28–31]. *EGFR* amplification has been proposed as an independent adverse prognostic factor in OAC and has been implicated in chemotherapy resistance [32]. Despite this strong biologic rationale, EGFR-directed therapies have shown limited clinical benefit in unselected patients with OAC. Trials of the tyrosine kinase inhibitor (TKI) Gefitinib [33] and anti-EGFR monoclonal antibody Cetuximab [34] failed to improve outcomes. Similarly, in REAL 3, the addition of monoclonal antibody panitumumab to epirubicin, oxaliplatin, and capecitabine (EOC) unexpectedly resulted in inferior survival. Subsequent analyses suggested that the negative result may have reflected antagonism between EGFR inhibition and anthracycline chemotherapy rather than a lack of biological relevance of EGFR itself. These findings emphasise the need for more refined biomarker-driven approaches and alternative combination strategies when targeting the EGFR pathway in OAC (NCT00824785) [35,36].

We observed divergent prognostic associations within the RTK signalling axis. While EGFR amplification was independently associated with poor survival, gain-of-function variants or amplifications in KRAS and ERBB2 showed non-significant trends towards improved survival. Previous large-scale genomic analyses have demonstrated that KRAS and ERBB2 alterations are mutually exclusive in OAC [6], suggesting that activation of the RTK pathway can occur through distinct molecular routes. Our findings further suggest that these alternative genomic events may define biologically and clinically distinct subgroups, highlighting that RTK pathways activation is not uniformly associated with prognosis.

Ryanodine receptor 1 (*RYR1*), involved in Ca2+ signalling, emerged as a recurrently altered gene associated with poor prognosis in our cohort. To our knowledge, *RYR1* has not previously been reported as a commonly altered gene or prognostic biomarker in OAC. *RYR1* alterations were more frequent in more advanced staged disease and were independently associated with poor survival. Consistent with observations in other tumour types, *RYR1* and especially *LRP1B* mutations were enriched among tumours with high TMB. Given their large genomic size, alterations in these genes are thought to arise through accumulation of passenger mutations in hypermutated tumours, rather than representing primary drivers [37]. Nonetheless, LRP1B and potentially RYR1 may have clinical utility as surrogate markers of a hypermutated phenotype that may benefit from immunotherapy.

Strengths of our study include the unique surgery-only cohort, in which all patients underwent a standardised *en bloc* oesophagectomy with extended lymphadenectomy potentially improving pathological lymph node staging, together with more than 11 years’ minimum follow-up. The genomic landscape observed was broadly consistent with previous reports, although we observed higher mutation frequencies for *LRP1B, RYR1, PIK3CA*, and *ARID2*. The principal limitation is the relatively small cohort size, and therefore the associations with survival require independent validation. Despite this limitation, our findings reproduced established adverse prognostic associations between *CCND1* amplification [13–15] and *EGFR* aberration [28–32] and overall survival. In addition, we identified several potentially novel prognostic biomarkers. *RYR1*, a calcium signalling gene not previously implicated in OAC prognosis, was recurrently altered and independently associated with poorer survival. We also found that aberrations in chromatin remodelling genes, particularly *KMT2D* and *ARID2*, were associated with worse survival for OAC, suggesting that epigenetic dysregulation contributes to aggressive tumour biology in OAC. If validated, these findings provide further rationale for investigating epigenetic modulation strategies in molecularly-selected patients.

## MATERIALS AND METHODS

### Study design, setting and participants

We conducted a retrospective cohort study of patients with a diagnosis of adenocarcinoma of the oesophagus or gastro-oesophageal junction (Siewert type I or II) and undergoing surgery with curative intent at Lund University Hospital, Sweden between September 2006 and October 2011. Additional inclusion criteria were that *en bloc* oesophagectomy with extended lymphadenectomy was performed and fresh frozen tissues were collected intra-operatively for research purposes. *En bloc* oesophagectomy was performed in the same manner for all patients by four senior upper gastrointestinal surgeons using a highly standardised method. Details of the operation have been described elsewhere [38,39].

Due to an administrative preference at the time, none of the patients, regardless of tumour stage, received neo-adjuvant or adjuvant non-operative chemo-, radio- or other non-operative therapy. Follow-up was undertaken by the treating surgeons up to 1st July 2022.

Preparatory laboratory studies were performed at the Centre for Applied Medical Research and next generation sequencing (NGS) was performed at the Kinghorn Cancer Centre, both at St Vincent’s Hospital, Sydney, Australia.

### Variables, data sources and measurement

We collected the following information on clinicopathological characteristics for each patient : age at operation, sex, cancer location recorded by pathologist (Siewert I (distal oesophagus), Siewert II (gastro-oesophageal junction)), tumour grade (well, moderately, poorly differentiated), pathology-defined TNM (pTNM) stage classified according to AJCC 8th edition, margin status (R0 no microscopic or macroscopic disease, R1 microscopic disease without macroscopic disease), total number of lymph nodes resected, number of lymph nodes positive for cancer, and lymph node ratio (number of positive lymph nodes: total lymph nodes resected).

Primary tumour tissues and matched normal squamous oesophagus tissues were collected from each individual for laboratory studies.

The primary outcome for genomic prognostic analyses was OAC-specific death. We also recorded all-cause death to assess overall survival by TNM stage. Date and cause of death were recorded from the Cause of Death Register at the Swedish National Board of Health and Welfare. Time to death was calculated from the date of surgery to the date of death, or censored at 1 July 2022 if no death was recorded.

### NGS library preparation and sequencing

Sequencing libraries were prepared using a targeted 451-cancer gene panel (Table 1 in McCabe et al. 2019) [40]. Briefly, DNA was extracted from fresh frozen tumours and matching normal squamous oesophageal tissues using Qiagen’s DNeasy kit (cat #69504). Library preparation was performed on 100ng of DNA using the KAPA HyperPlus Kit (Roche, cat #KK8514) with selection against 2.01Mb target regions. Prepared libraries were sequenced 2x150bp on HiSeq2500 at target coverage of 400x to yield ∼10M reads.

### Bioinformatic and statistical analysis

Raw fastq files were processed using the bioinformatics pipeline as described in McCabe et al. (2019) [40]. Alignment to b37 genome was performed using BWA-MEM [41]. The aligned files were sorted using Novosort (http://www.novocraft.com). Picard metrics were used for quality check (http://broadinstitute.github.io/picard). Samples that failed our minimal quality requirements (total reads < 5M, Percent Unique reads aligned <95%, Target Median Coverage <100X) were excluded from analysis.

Consensus somatic variant calling was performed using a combination of Strelka2 [42], VarDict [43] and Mutect2 [44], whereby reported somatic variants must be called using at least two of the aforementioned callers. Single nucleotide variant (SNV) annotation was performed using Ensembl Variant Effect Predictor (VEP) [45]. Copy number changes were called using CNVkit [46]. The purity and ploidy estimations were calculated using PureCN [47]. Driver mutations were inferred using a combination of MutPanning [48], oncodriveFML [49] and annotation with OncoKB [50].

We summarised the clinical and tumour characteristics of the study cohort as percentages or medians and range and visually displayed these characteristics for each patient with their survival time using bar plots ordered by survival time. We evaluated the strength of association between established prognostic factors (age, TNM stage, tumour grade, cancer location, lymph node ratio), and overall survival in the study cohort by estimating unadjusted and adjusted hazard ratios (HR) and 95% confidence intervals (CI) using Cox proportional hazards models and presented the results in forest plots. We included age, sex, TNM stage and cancer location in these multivariable models. We did not include tumour grade and lymph node ratio in the models due to their strong correlation with TNM stage.

We compared the frequency of the 25 most frequently mutated genes in our cohort with the frequencies reported for OAC in The Cancer Genome Atlas (TCGA) [7], Memorial Sloan Kettering Cancer Centre (MSKCC) [8] and the Dana-Faber Cancer Institute/Broad Institute (DFCI/BROAD) [5] datasets. We used the independent proportions z-test to test the null hypothesis that gene frequencies would not vary between OAC populations.

Additionally, we computed the frequency of copy number gains and losses across our cohort, using a log_2_ fold change of ±0.3 to designate a gain or loss. Significantly enriched loci were detected using GISTIC v2.0 with qval <0.1 [51]. The overall frequency of gains, losses and mutations for commonly aberrated cancer pathways were visualised using PathwayMapper [52].

We evaluated the association between each of the most commonly aberrated genes and OAC-specific survival using multivariable Cox proportional models to estimate hazard ratios adjusted for age and stage. We visually displayed the frequency of SNVs and CNVs grouped by impacted pathway across the patient cohort stratified by tumour stage as a preliminary assessment of whether specific variants were more likely to be present in tumours diagnosed at an early or late stage.

We assessed tumour mutational burden for each patient tumour sample, calculated as the number of coding mutations per MB of coding regions.

### Ethics approval

Institutional ethics approval and material transfer agreement was obtained from Skane University Hospital, Lund, Sweden (2014/224) and from St. Vincent’s Hospital, Sydney, Australia (ETH03452). Informed consent was obtained from all participating individuals. All methods were carried out in accordance with relevant guidelines and regulations.

## Acknowledgements

This research was funded by Kinghorn foundation (A.T., M.N., L.G.), Bioplatforms Australia (A.J.), National Health and Medical Research Council (NHMRC (T.P.U., S.J.L., A.J.L-M., Y.J.L., D.P., M.J.B., J.R., R.V.L.)) Ideas Grant 2012513, St. Vincent’s Clinic Foundation (R.V.L.).

## Author contribution

D.F., A.J.L-M., and R.V.L. were involved in the conception and design of the study. A.T., T.P.U., S.J.L., D.F., A.J.L-M., J.J. and R.V.L. carried out the acquisition of data and/or analysis and interpretation of data. The manuscript was written by A.T., T.P.U., S.J.L., M.B., and R.V.L. All authors were involved in review of the manuscript and had final approval of the submitted and published versions.

## Data availability statement

Data has been deposited at the European Genome-phenome Archive (EGA), which is hosted by the EBI and the CRG, under accession number EGAS50000002190 (https://ega-archive.org/studies/EGAS50000002190). Further information about EGA can be found at https://ega-archive.org and The European Genome-phenome Archive in 2021: 10.1093/nar/gkab1059. Analysis code and processed data are available at github.com/trinhan/EAC_Lund_En-bloc. Individual patient data are not available.

