## supplementary_figure1 for "Genomic correlates with survival after treatment of oesophageal adenocarcinoma by surgery alone"

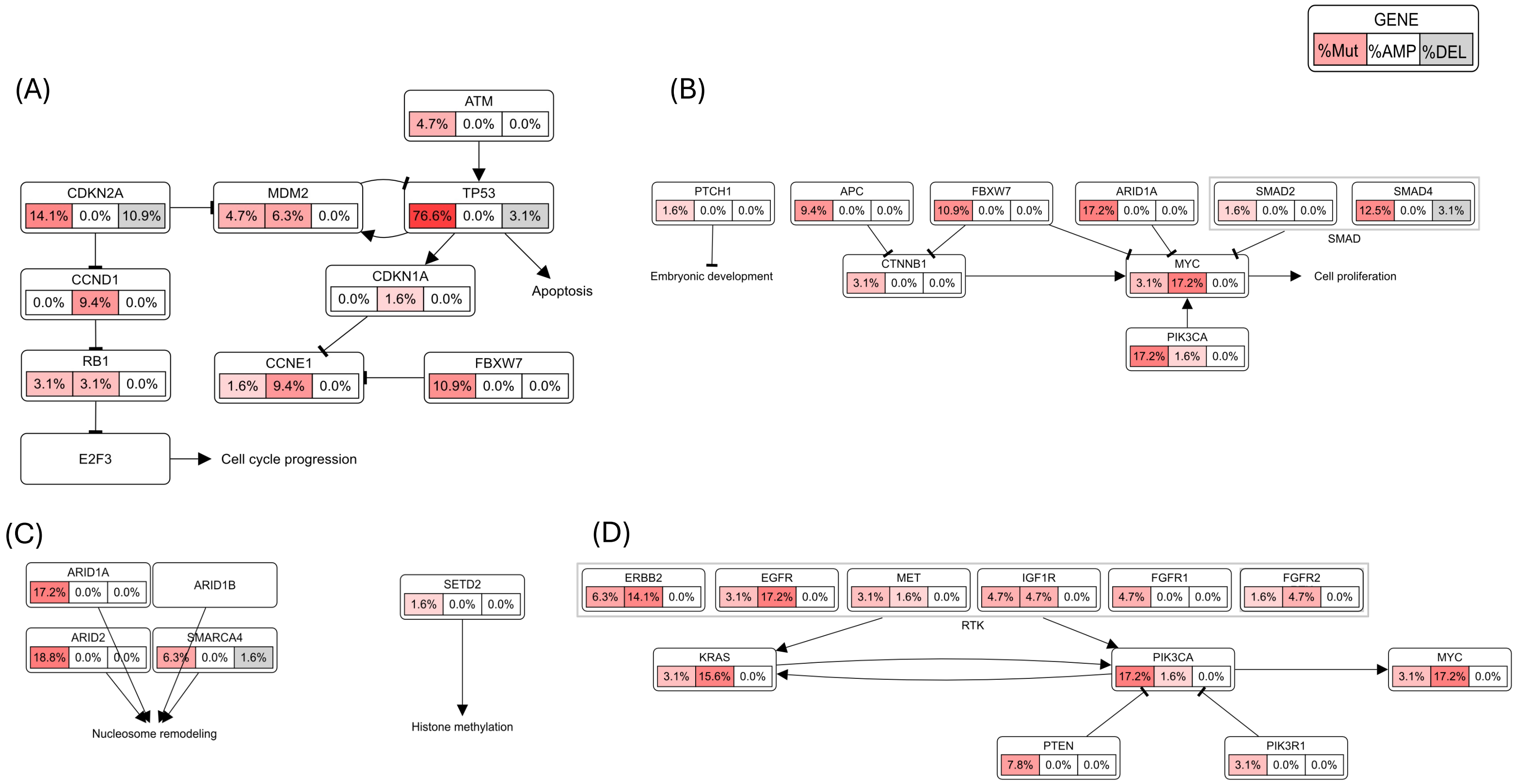


**Supplementary Figure 1:** Frequency of variants in genes associated with **A.** cell-cycle signalling, **B.** Wnt pathway, **C.** histone remodelling **D.** RTK signalling across the cohort, as visualised using pathway mapper.  For each gene, left cell shows mutation frequency (%), middle cell shows amplification (% gain) and right cell deletion (% loss).
